# Relationship Between Patient Sex and Serum Tumor Necrosis Factor Antagonist Drug and Anti-Drug Antibody Concentrations in Inflammatory Bowel Disease; A Nationwide Cohort Study

**DOI:** 10.1101/2021.06.01.21258121

**Authors:** Mohammad Shehab, Hajer Alasfour, Israa Abdullah, Ghadeer Alhendi, Anwar Alhadab, Ahmad Alfadhli, Ali H. Ziyab, Robert Battat

## Abstract

**Background:** Anti-drug antibodies to infliximab (ATI) and adalimumab (ATA) are associated with loss of response to tumor necrosis factor antagonist (anti-TNF) therapy in inflammatory bowel disease (IBD). We evaluated the relationship between patient sex and serum TNF antagonist drug and antibody concentrations in inflammatory bowel disease

**Methods:** A nationwide multicenter retrospective cohort study was conducted by evaluating patients’ charts from July 2018 until September 2021. The effect of patient sex on anti-drug antibodies and serum drug concentration in patients with IBD across 7 hospitals was investigated. A subgroup analysis also investigated the effect of anti-TNF combination therapy. Geometric means were calculated, and multiple linear regression was used to estimate the adjusted ratio of geometric means (RoGM) and their 95% confidence intervals (CI).

**Results:** In the total study sample (n = 1093), males receiving infliximab had higher anti-drug antibody concentrations (38.3 vs. 22.3 AU/ml; aRoGM = 1.72, 95% CI: 1.30-2.27, p-value <0.001) compared to females. Additionally, infliximab serum drug concentrations among males were lower compared to females (2.6 vs. 4.1 ug/ml; aRoGM = 0.62, 95% CI: 0.44-0.88, p-value = 0.007). In the subgroup analysis (n = 359), male compared to female patients on combination therapy with infliximab and immunomodulators had similar anti-drug antibody concentrations (30.2 vs. 21.9 AU/ml; aRoGM = 1.38, 95% CI: 0.79-2.40, p-value = 0.254). There was no difference in the anti-drug antibody and serum drug concentrations among males and females on adalimumab.

**Conclusion:** In patients receiving infliximab, anti-drug antibodies were higher in males than females. Consistent with this, serum drug concentrations were lower in males than females on infliximab. There was no difference in anti-drug antibody and serum drug concentrations among males and females on adalimumab. In addition, no difference in anti-drug antibodies between males and females receiving anti-TNF combination therapy was observed.

## Introduction

Tumor necrosis factor antagonist (anti-TNF) therapies are commonly used for the management of moderate to severe inflammatory bowel disease (IBD).^1-4^ However, about one-third of patients treated with anti-TNF therapy develop primary treatment failure (primary non-response), in which a lack of response is observed in induction therapy.^5,6^ Furthermore, approximately half of patients with initial response may experience secondary loss of response by losing treatment effect during the maintenance of remission.^7^ One of the most common causes of treatment failure is immunogenicity, the formation of anti-drug antibodies, which is also associated with low or undetectable drug serum concentrations.^8,9^ The serum drug concentrations of anti-TNF therapy might also vary depending on the severity of the disease, degree of inflammation, concurrent use of immunomodulator, patient sex, serum albumin concentration, body mass index (BMI), and genetic factors.^7,8^

Previous data show that anti-drug antibodies exist in over 20% of IBD patients treated with anti-TNF therapy.^10^ Additionally, patient sex and body weight significantly influence the pharmacokinetics of infliximab as its clearance has been shown to be increased in the presence of anti-drug antibodies and low serum albumin.^11^ On the other hand, combination therapy, the concurrent administration of an immunomodulator with an anti-TNF, has been associated with improvement in pharmacokinetics by decreasing immunogenicity and increasing serum drug concentrations.^1-4^ With respect to infliximab, the SONIC and UC-SUCCESS trials demonstrated that the use of infliximab combination therapy is superior to monotherapy in reducing immunogenicity and maintaining remission.^12,13^ Conversely, DIAMOND trial and two other meta-analyses, by Kopylov et al. and Chalhoub et al., demonstrated that adalimumab combination therapy is associated with limited impact on maintenance of clinical remission or response.^3,14-16^ When considering anti-TNF therapy for pediatric patients with IBD, the ECCO-ESPGHAN guidelines recommend the use of infliximab combination therapy, with an immunomodulator, to reduce the risk of developing anti-drug antibodies to infliximab (ATI). However, for adalimumab, it is preferred to be prescribed as a monotherapy when started as the first anti-TNF agent in children.^17,18^ When combination therapy is used in pediatric patients, it is recommended to stop the concomitant use of the immunomodulator after 6–12 months, and the benefits of continuing combination therapy should be weighed against the risk of adverse events.^17^

A large-scale real world studies with patient-level data are lacking on the relationship between patient sex and anti-TNF drug and anti-drug antibody concentrations. Additionally, the impact of combination therapy on anti-TNF pharmacokinetics when accounting for sex has not been described. To address these knowledge gaps, this study utilized a large cohort with patient level data to determine the relationship between patient sex and immunogenicity of infliximab and adalimumab when accounting for important factors such as albumin and concomitant immunomodulator use.

## Materials and Methods

### Study design

A nationwide multicenter retrospective cohort study was conducted to measure the effect of patient sex on anti-TNF anti-drug antibodies and serum drug concentrations in patients with inflammatory bowel disease (IBD). A subgroup analysis was performed at an inflammatory bowel disease center for patients who received either infliximab or adalimumab monotherapy or in combination with an immunomodulator. This study was performed and reported in accordance with Strengthening the Reporting of Observational Studies in Epidemiology (STROBE) guidelines.^19^ The study protocol was reviewed and approved by the standing committee for coordination of health and medical research (IRB 2020/1410).

We included patients that: (1) were previously diagnosed with inflammatory bowel disease (2) had been tested for an anti-drug antibody and/or serum drug trough concentrations reactively or proactively and (3) were receiving an anti-TNF therapy for at least 6 weeks at the time of measurement. (4) who received regular standard dose anti-TNF therapy. Patients who had past medical history of other autoimmune diseases, such as inflammatory arthritis, or were on immunosuppressant therapies for other non IBD medical conditions were excluded. The data were collected from patients’ electronic medical records from seven different hospitals. Data were collected from July 22nd, 2018 until September 1st, 2021. A subgroup analysis, from the total sample, was performed at an inflammatory bowel disease center for patients who received either infliximab or adalimumab combination or monotherapy.

Patient’s characteristics were obtained from patient’s electronic medical records including age, body mass index (BMI), type and extent of IBD at the time of serum drug/antibody concentration measurements.

Diagnosis of inflammatory bowel disease (IBD) was made according to the international classification of diseases (ICD-10 version:2016). Patients were considered to have IBD when they had ICD-10 K50, K50.1, K50.8, K50.9 corresponding to Crohn’s disease (CD) and ICD-10 K51, k51.0, k51.2, k51.3, k51.5, k51.8, k51.9 corresponding to ulcerative colitis (UC).^20^

## Study Definitions

Patients were considered to have active inflammation if they have one of the following within 14 days from serum drug/antibody concentration measurements: (1) C-reactive protein (CRP) levels above 10 mg/L or (2) stool fecal calprotectin (Fcal) more than 250 ug/g or (3) receiving steroids. Patients were considered to be receiving steroids if they were concomitantly receiving budesonide, methylprednisolone, hydrocortisone, prednisone/prednisolone, or any steroidal agent. Moreover, patients who received an immunomodulator (such as azathioprine, 6-mercaptopurine or methotrexate) concurrently with an anti-TNF therapy were classified to be on combination therapy while patients on infliximab or adalimumab alone were classified to be on monotherapy.

Anti-drug antibody and serum drug concentration samples from all the hospitals were measured at one central immunology laboratory. Anti-drug antibodies were considered detectable at levels >5 AU/ml for infliximab or >10 AU/ml for adalimumab. Additionally, serum drug concentrations/antibody levels were collected only at trough levels, i.e before the next scheduled dose. A serum drug level of ≥5 ug/ml, and ≥7.5 ug/ml was considered therapeutic for infliximab and adalimumab, respectively. Trough serum drug and antidrug antibody concentrations were performed either reactively, e.g. due to treatment failure, or proactively to optimize therapy, e.g at week 14 for infliximab, as per each physician clinical judgment and practice. Anti-drug antibodies and serum drug concentrations were analyzed as continuous variables while applying log-transformation to account for skewness in the data and maximize the value of the continuous measurements (see discussion).

### Outcomes

The primary outcome was to compare anti-drug antibody levels between male and female patients on infliximab and adalimumab. In addition, the association between patient sex and serum drug concentration for infliximab and adalimumab was evaluated.

Secondary outcomes, in sub-analyses, evaluated the impact of combination therapy among male and female patients on infliximab or adalimumab anti-drug antibody levels. The association between serum drug concentration and combination therapy in both sexes was estimated as well. Moreover, combination therapy was compared to monotherapy in terms of anti-drug antibody levels and serum drug concentration.

### Statistical analysis

Analyses were conducted using SAS 9.4 (SAS Institute, Cary, NC, USA). The statistical significance level was set to α = 0.05 for all association analyses. Descriptive analyses were conducted to calculate frequencies and proportions of categorical variables in the total study sample (n = 1093) and subsample, which was part of the main total sample, (n = 359). To account for skewed distribution of the continuous variables (i.e., serum drug concentrations, and anti-drug antibodies), geometric means were estimated by log_10_-transformation of the data and subsequently taking the antilog of the calculated means on the transformed scale.^21^ Correlations between anti-drug antibody and serum drug concentration according to anti-TNF drug type (infliximab and adalimumab) were assessed using Spearman correlation coefficients (r). In the total study sample analysis, the effect of patient sex on anti-drug antibody and serum drug concentrations of both adalimumab and infliximab was assessed. In the subsample analysis, the effect of patient sex on infliximab and adalimumab use in combination or alone on anti-drug antibody levels and serum drug concentrations was assessed.

Associations of adalimumab and infliximab with log_10_-transformed anti-drug antibody and serum drug concentration (outcome variables) were evaluated using multiple linear regression models. Associations were assessed in the total sample and stratified by sex. In the total sample analysis, associations were adjusted for the effects of sex, age at assessment, and active inflammation. In the subgroup analysis, associations were adjusted for sex, age at assessment, and active inflammation. In the sex stratified analysis, associations were adjusted for the effects of active inflammation and age at assessment. Given that we regressed log_10_-transformed anti-drug antibody and serum drug concentration, taking the antilog of the linear regression coefficients (β) yields an adjusted ratio of geometric means (aRoGM), not the difference between geometric means. Hence, the related 95% confidence intervals (CIs) represent limits for RoGM with a null value of “1”.

Additional analyses were conducted to determine whether albumin can be a confounder of the assessed associations. Albumin was categorized as normal (≥40 g/L) and abnormal (<40 g/L). The association of albumin (normal vs. abnormal) with anti-drug antibodies to infliximab and adalimumab was assessed by applying the Wilcoxon rank sum test. This analysis allowed us to determine whether concentrations of anti-drug antibodies differ across albumin categories. Moreover, albumin was added to the multiple linear regression models to determine if confounding is present.

## Results

### Demographics

In total, 1093 patients (567 [51.9%] males) were included in the total study sample analysis and a total of 359 patients (187 [52.1%] males) were included in the subsample analysis. The total study sample and the subsample were similar in all characteristics investigated. Of the total study sample, 42.2% of patients were on infliximab and 57.8% were on adalimumab. Similarly, in the subsample, 40.9% and 59.1% of patients were on infliximab and adalimumab, respectively (Table 1). Among patients in the subsample, 62.9% used anti-TNF combination therapy. Specifically, of the 147 patients on infliximab, 114 (77.6%) were on combination therapy while among the 212 patients on adalimumab, 112 (52.8%) were on combination therapy.

**Table 1.**
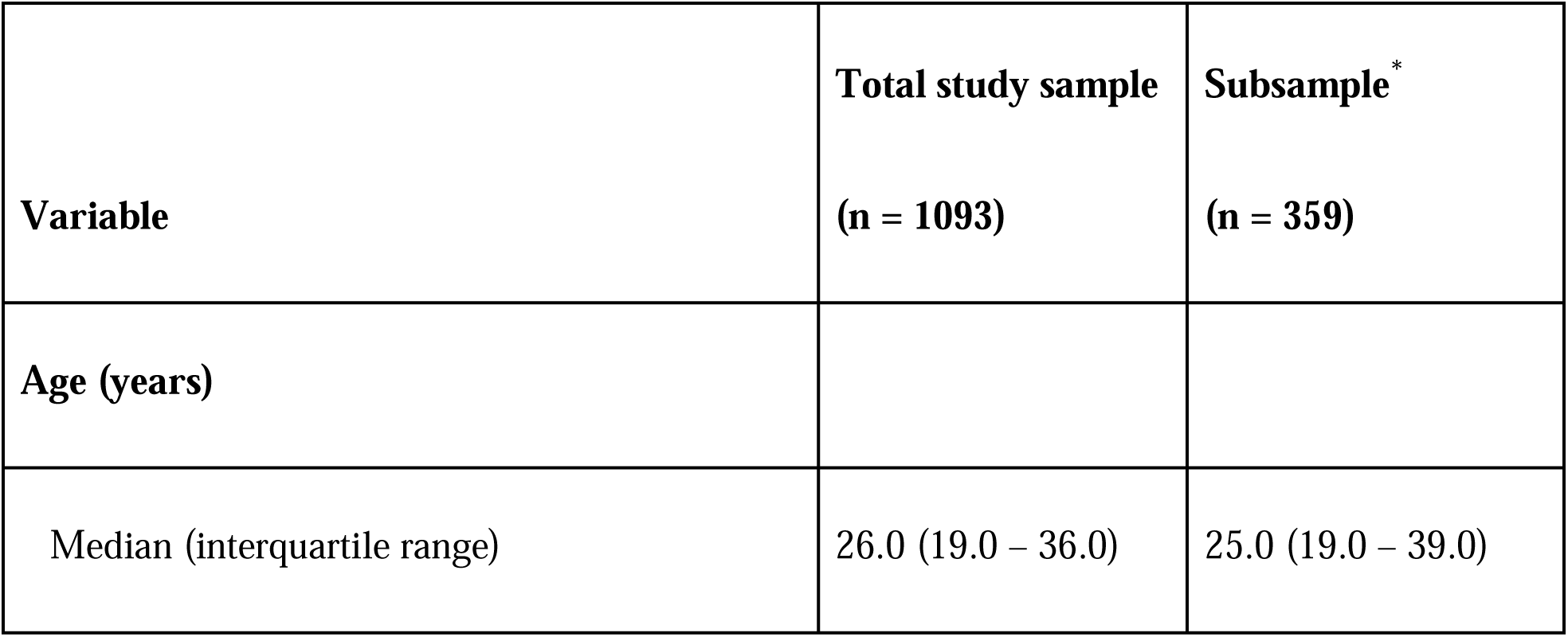
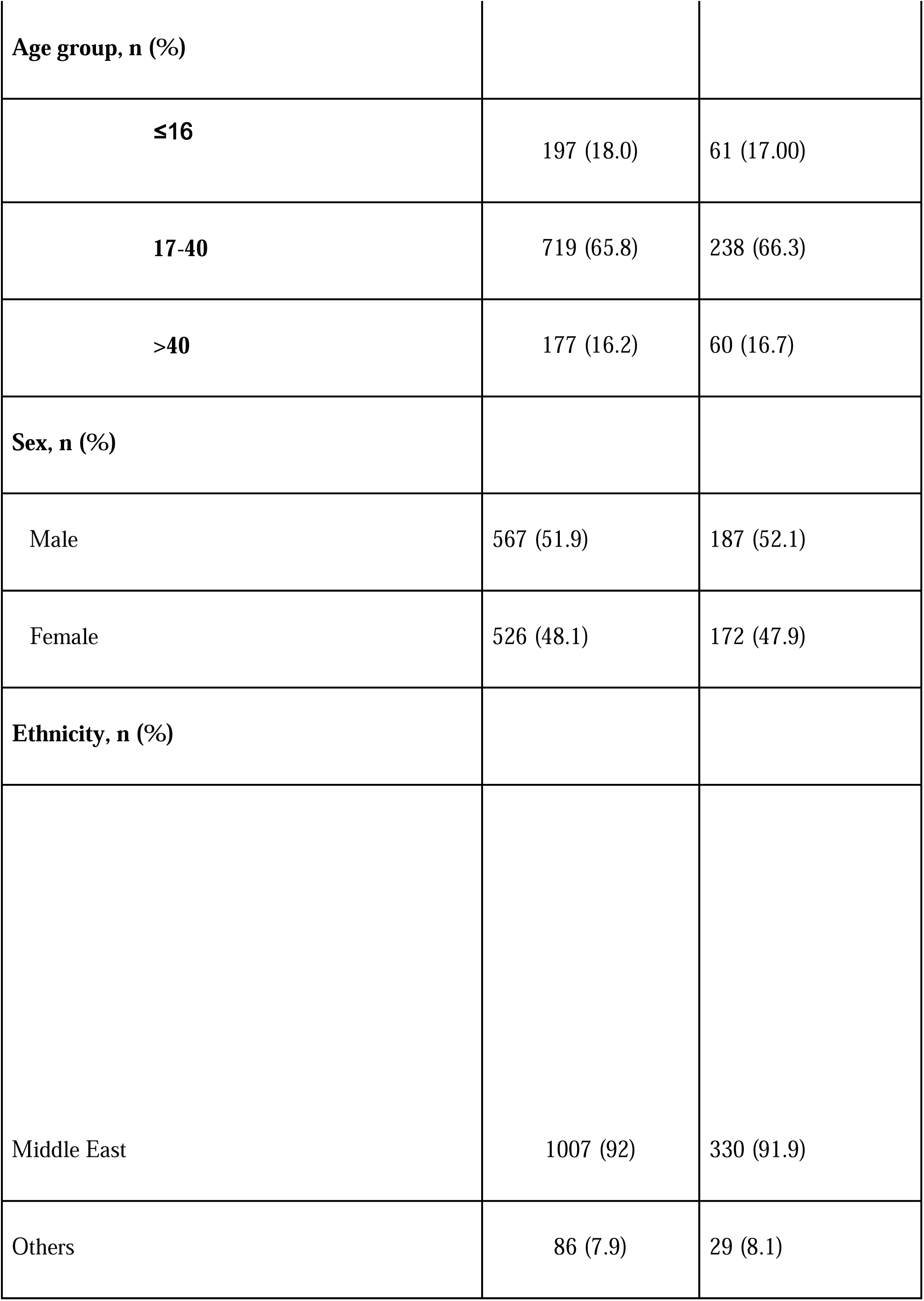
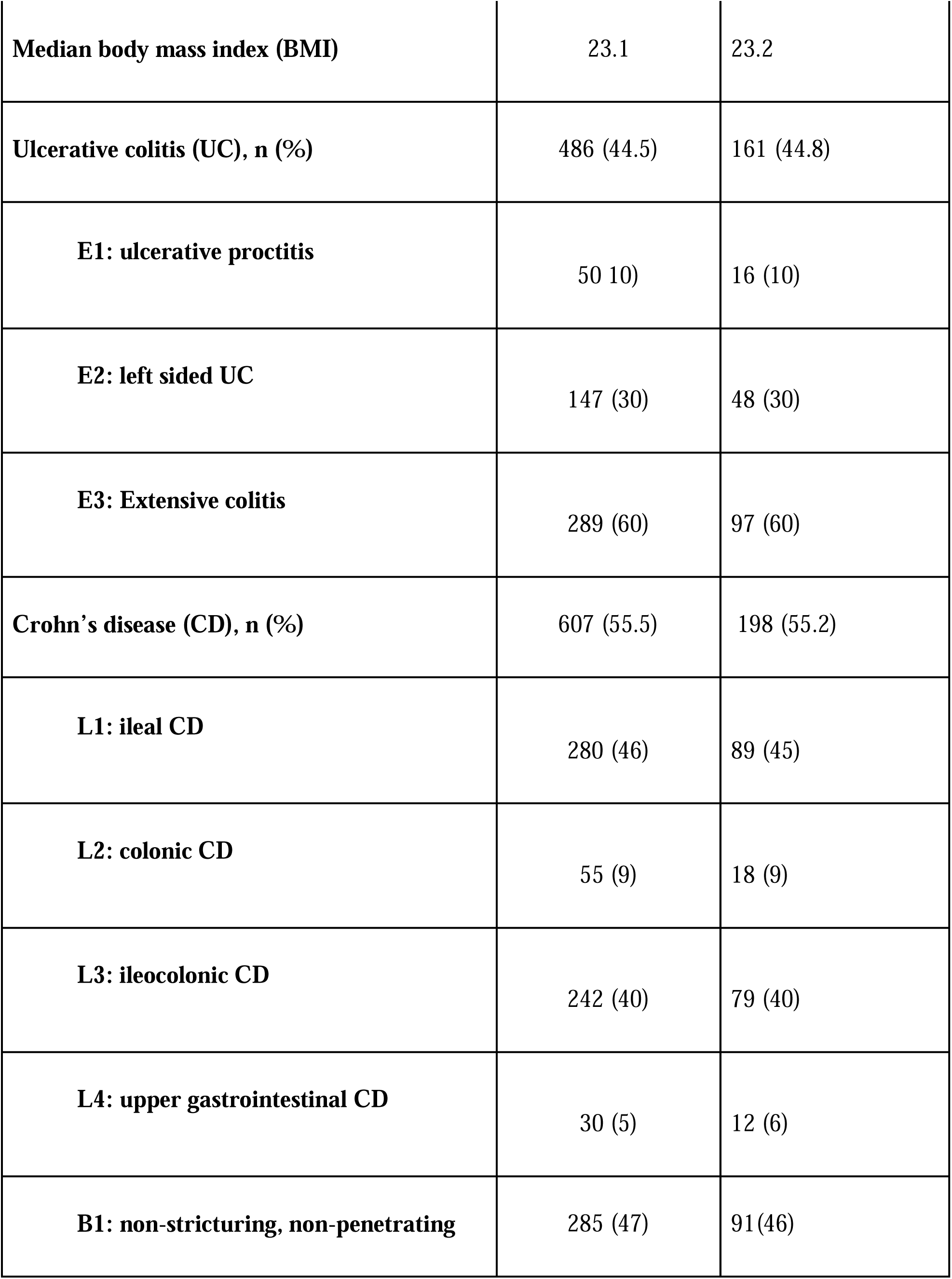
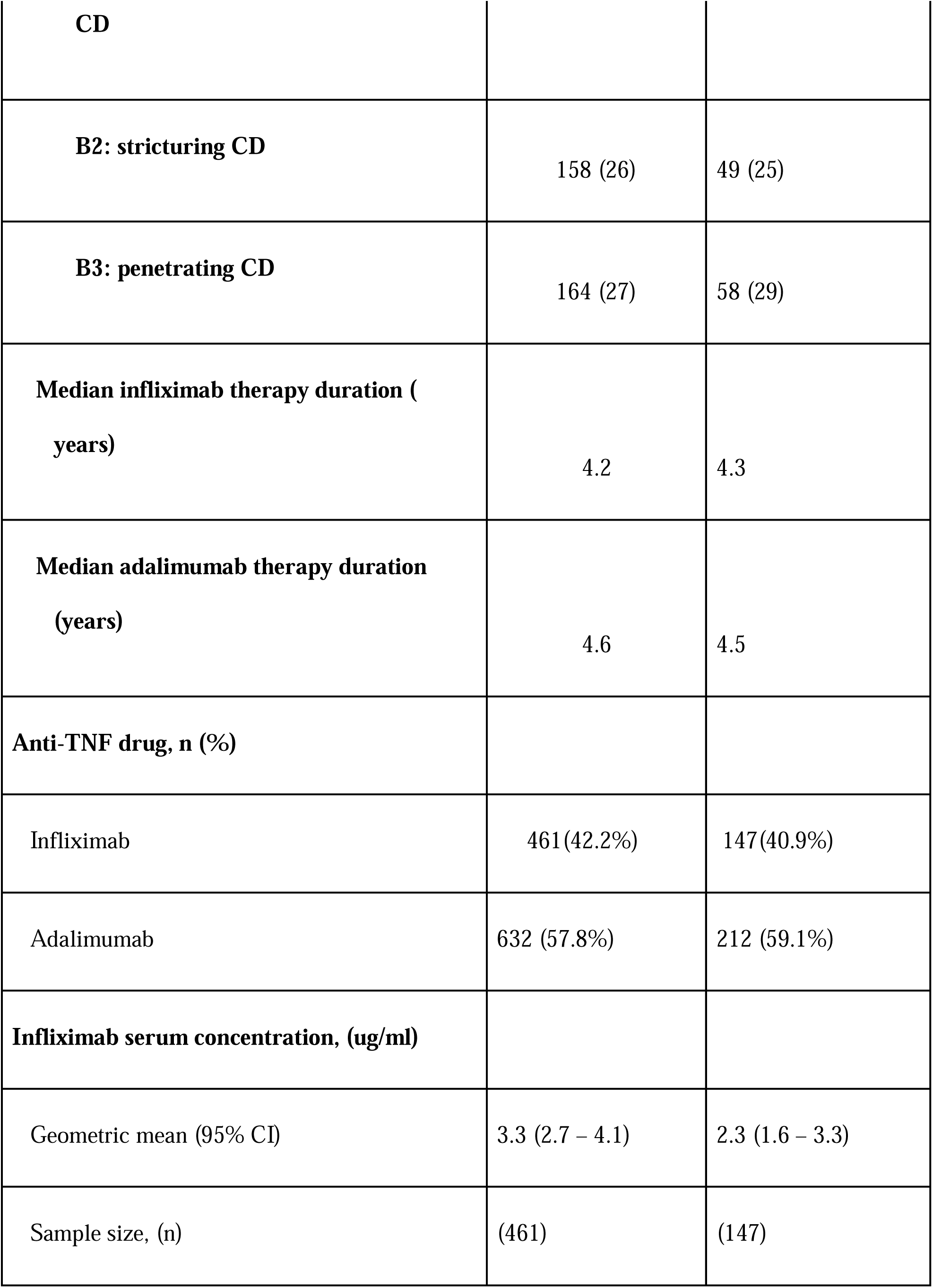
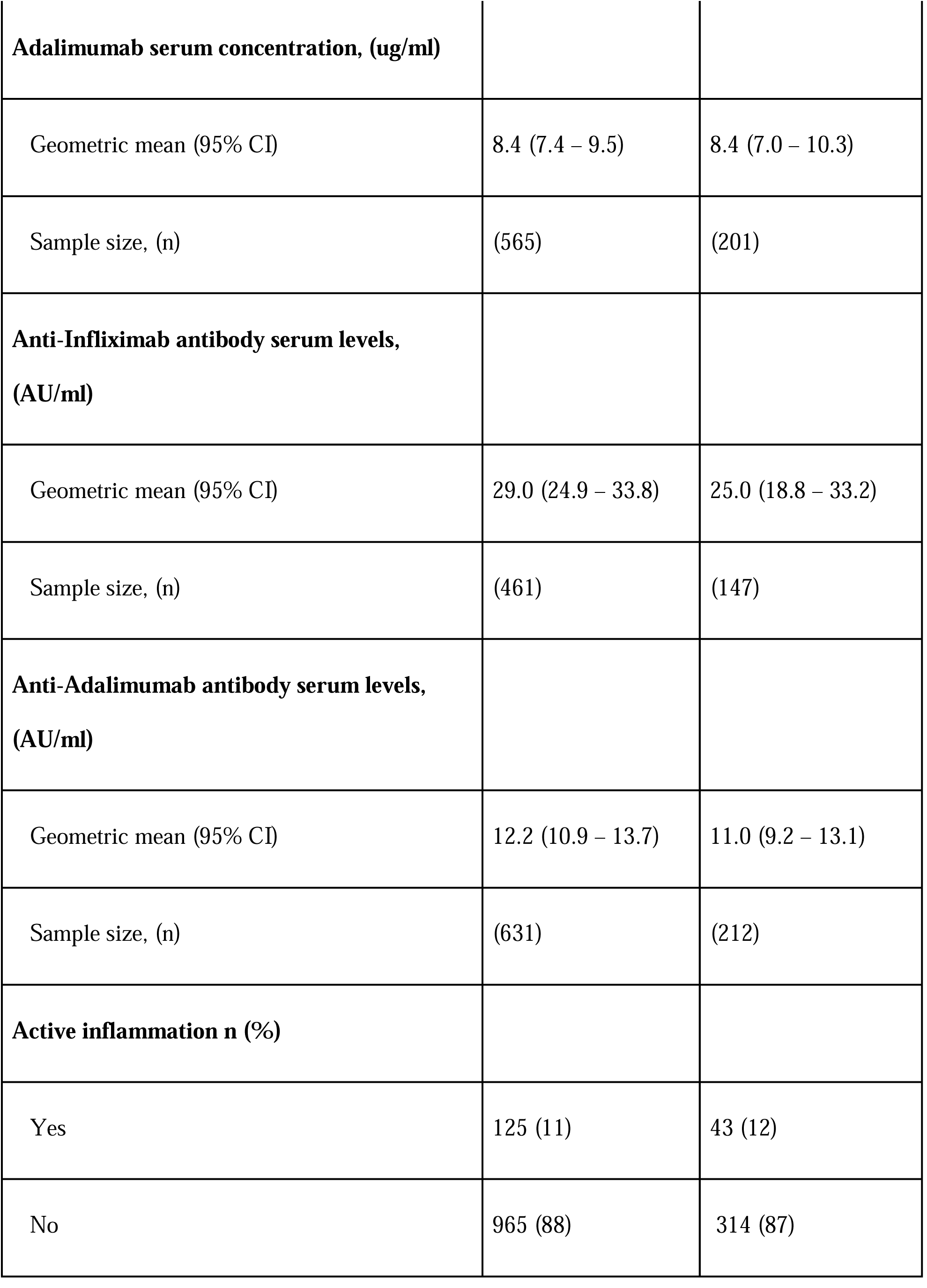
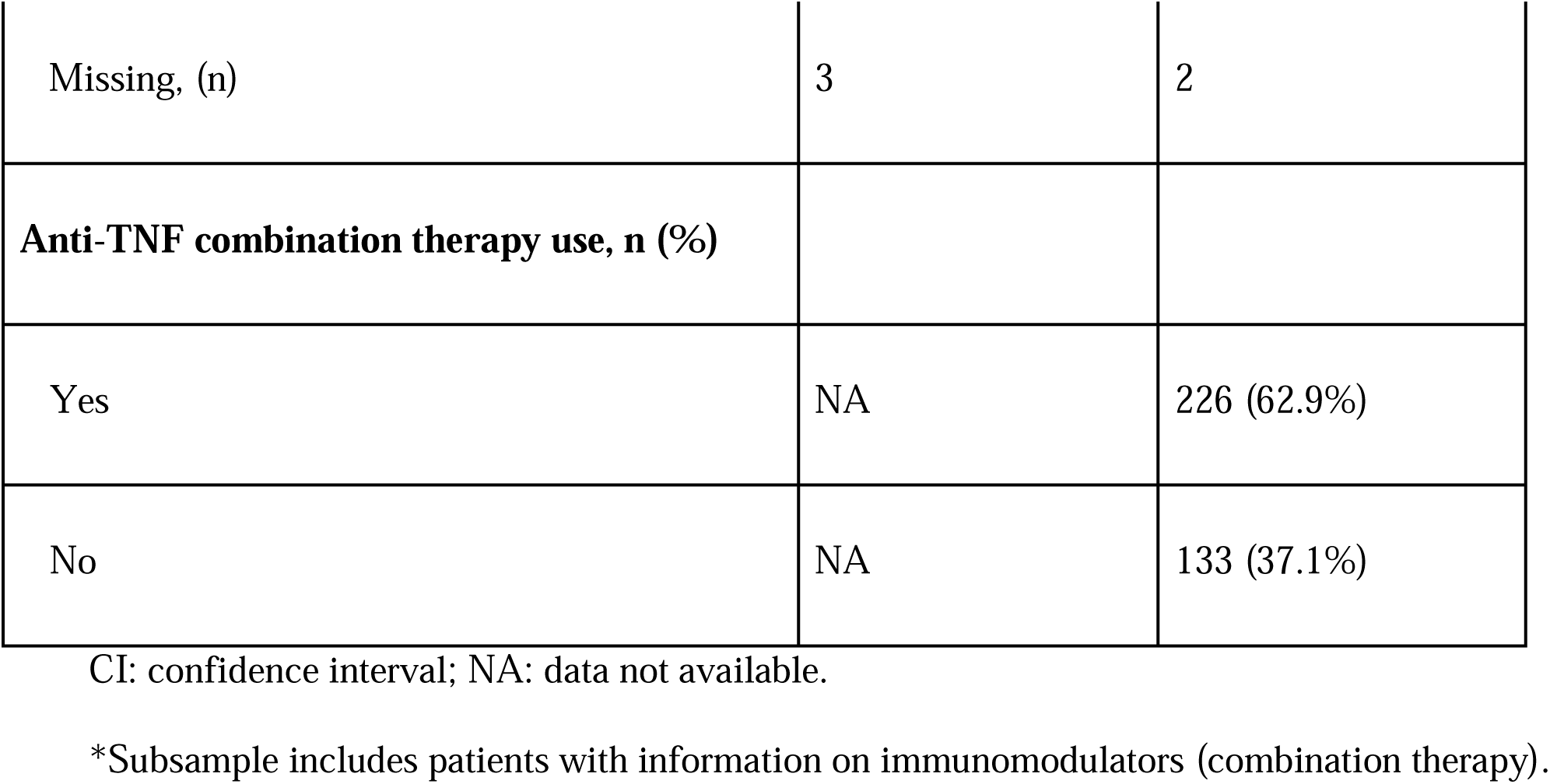
Characteristics of total study sample and subsample with information on combination therapy use

### Outcomes

Inter-sex comparisons of anti-drug antibody within anti-TNF therapy are shown in Table 2. Whilst the proportion of male and female patients are similar, male patients on infliximab had higher anti-drug antibody concentrations than female patients in the total sample (38.3 vs. 22.3 AU/ml; aRoGM = 1.72, 95% CI: 1.30-2.27, p-value <0.001) (Table 2, Figure 1a) and in the subsample sample (106.5 vs. 24.2 AU/ml; aRoGM = 4.39, 95% CI: 1.58-12.25, p-value =0.005) (Table 2, figure 1b). However, male and female patients on infliximab combination therapy in the subsample had similar anti-drug antibody levels (30.2 vs. 21.9 AU/ml; aRoGM = 1.38, 95% CI: 0.79-2.40, p-value = 0.254) (Table 2, Figure 1b).

**Table 2.**
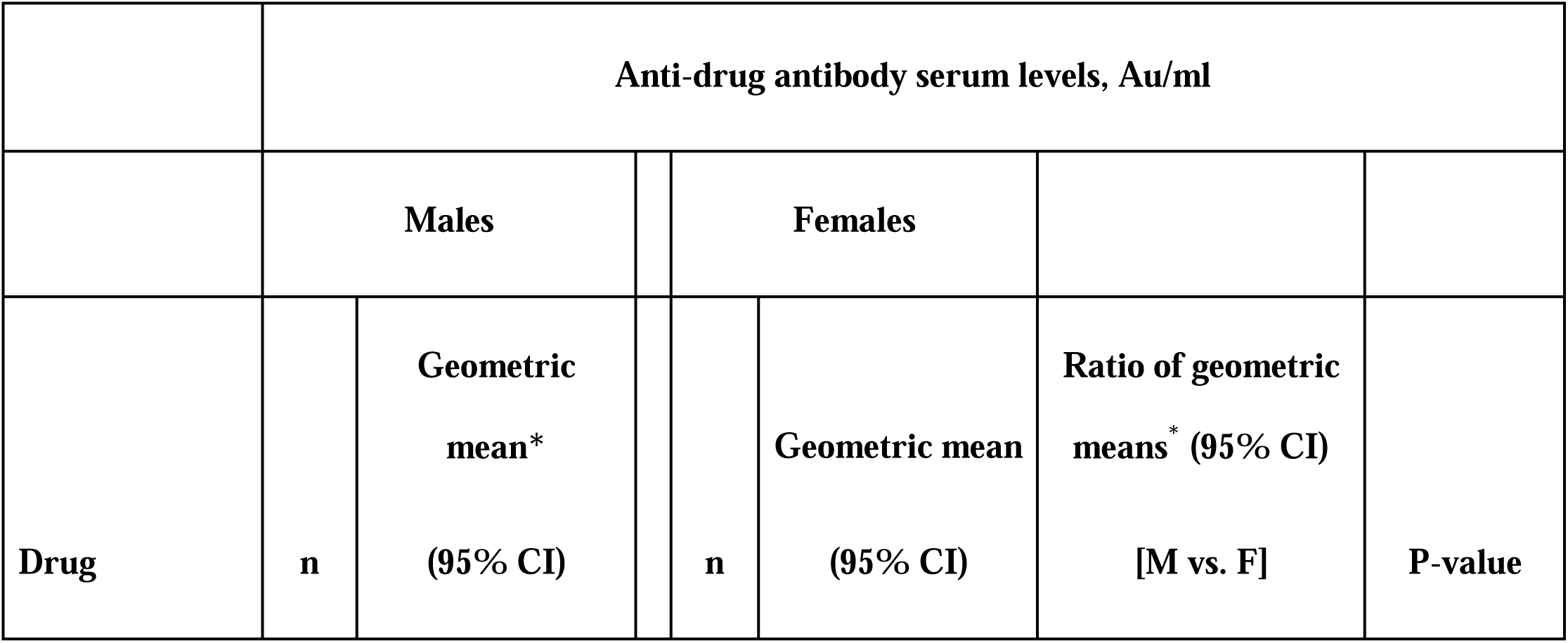
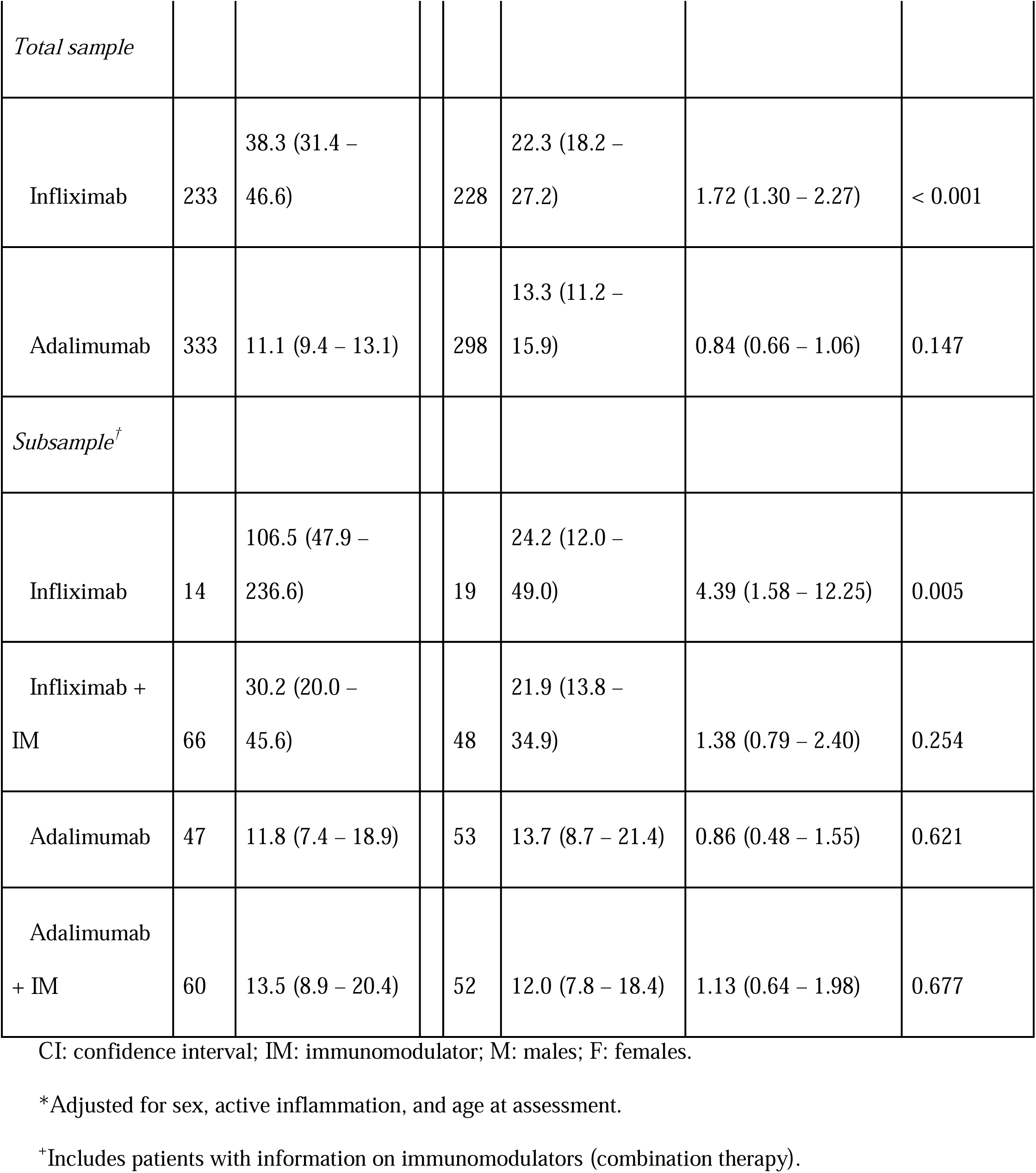
Anti-drug antibody serum levels stratified by sex according to monotherapy and combination therapy for infliximab and adalimumab use: inter-sex comparisons.

**Figure 1a.**
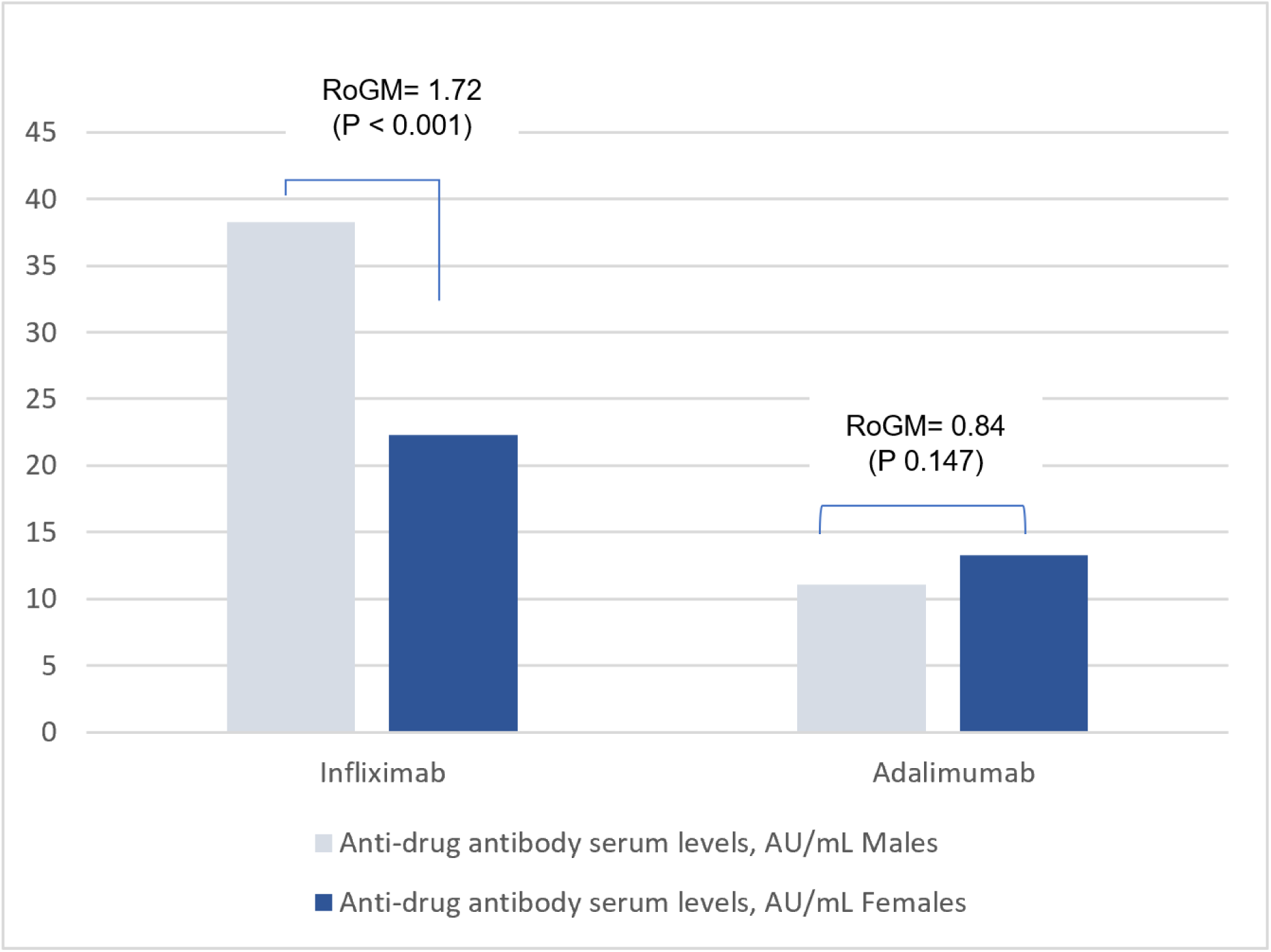
Anti-drug antibody levels in male and female patients with IBD in the total sample analysis for infliximab and adalimumab. The Y-axis corresponds to the geometric mean of the anti-drug antibodies serum levels.

**Figure 1b.**
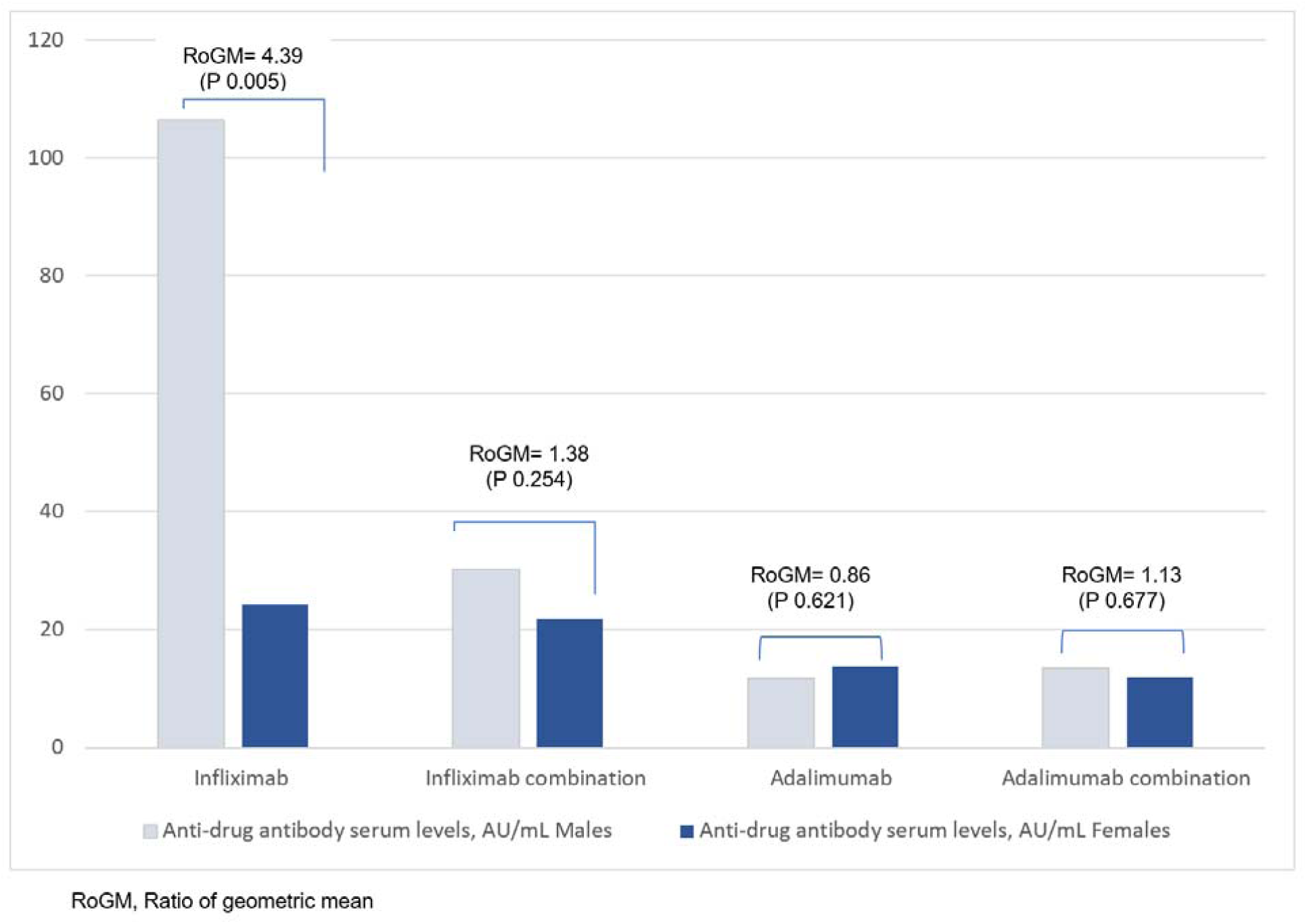
Anti-drug antibody serum levels in males and females with IBD in the subgroup analysis for Infliximab and Adalimumab. The Y-axis corresponds to the geometric mean of the anti-drug antibodies serum levels.

In the subsample, male and female patients on adalimumab alone had similar anti-drug antibody levels (11.8 vs 13.7 AU/ml; aRoGM=0.86, 95% CI: 0.48-1.55, p-value=0.62). Males and females on adalimumab combination therapy had similar anti-drug antibody levels as well (13.5 vs 12 AU/ml; aRoM=1.13, 95% CI: 0.64-1.98, p-value=0.67, Table 2, Figure 1b).

Associations between anti-TNF therapy and serum drug concentrations are shown in Table 3. In the total study sample, male patients had lower infliximab serum drug concentrations compared to female patients (2.6 vs. 4.1 ug/ml; aRoGM = 0.62, 95% CI: 0.44-0.88, p-value = 0.007). No other between sex differences in serum drug concentrations was observed (Table 3). Serum drug concentrations were similar among patients on infliximab alone compared to those on infliximab combination therapy (Supplementary Table s1). Similarly, no difference in serum drug concentrations was observed among patients on adalimumab alone and those on adalimumab combination therapy (Supplementary Table s1).

**Table 3.**
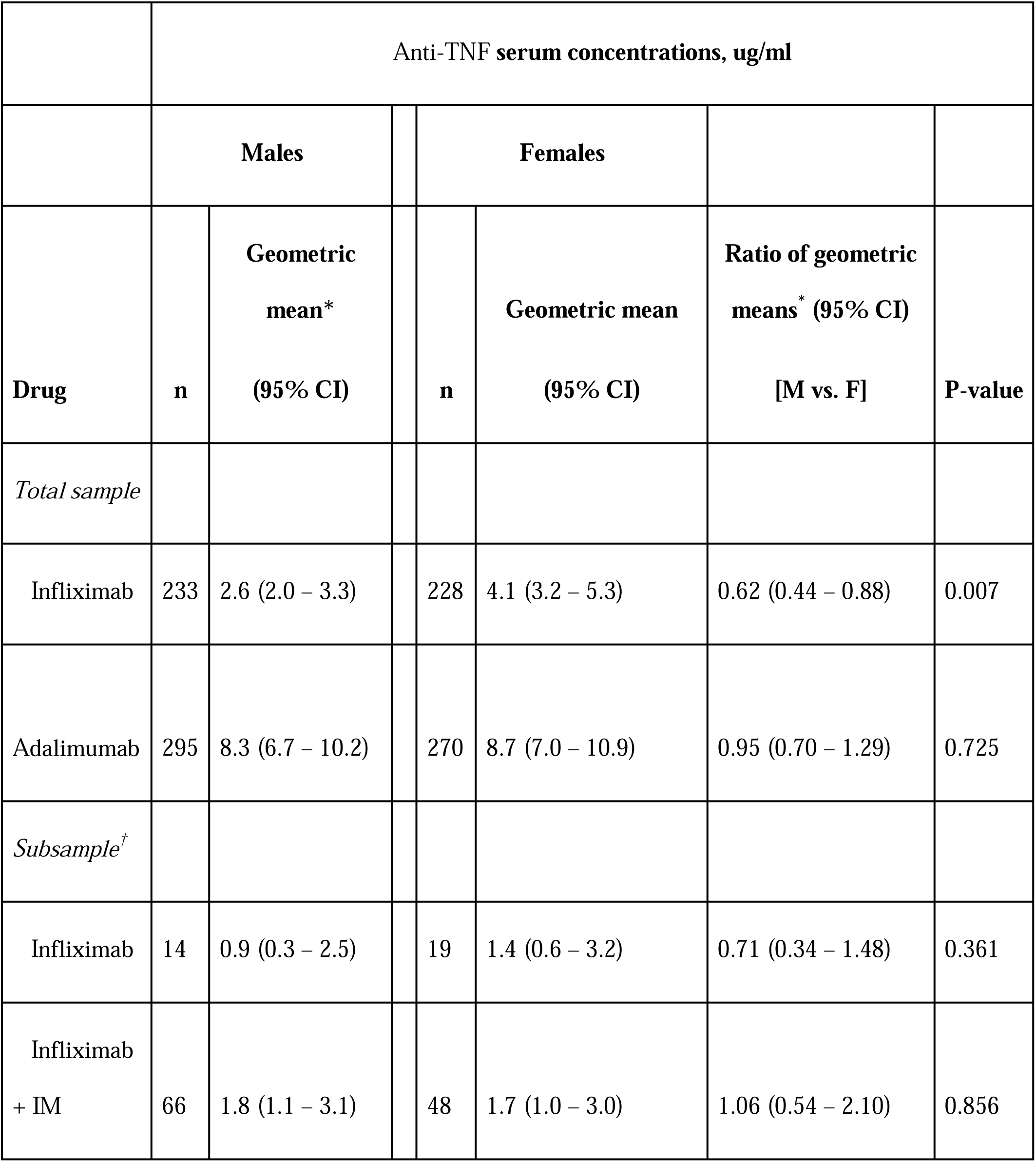
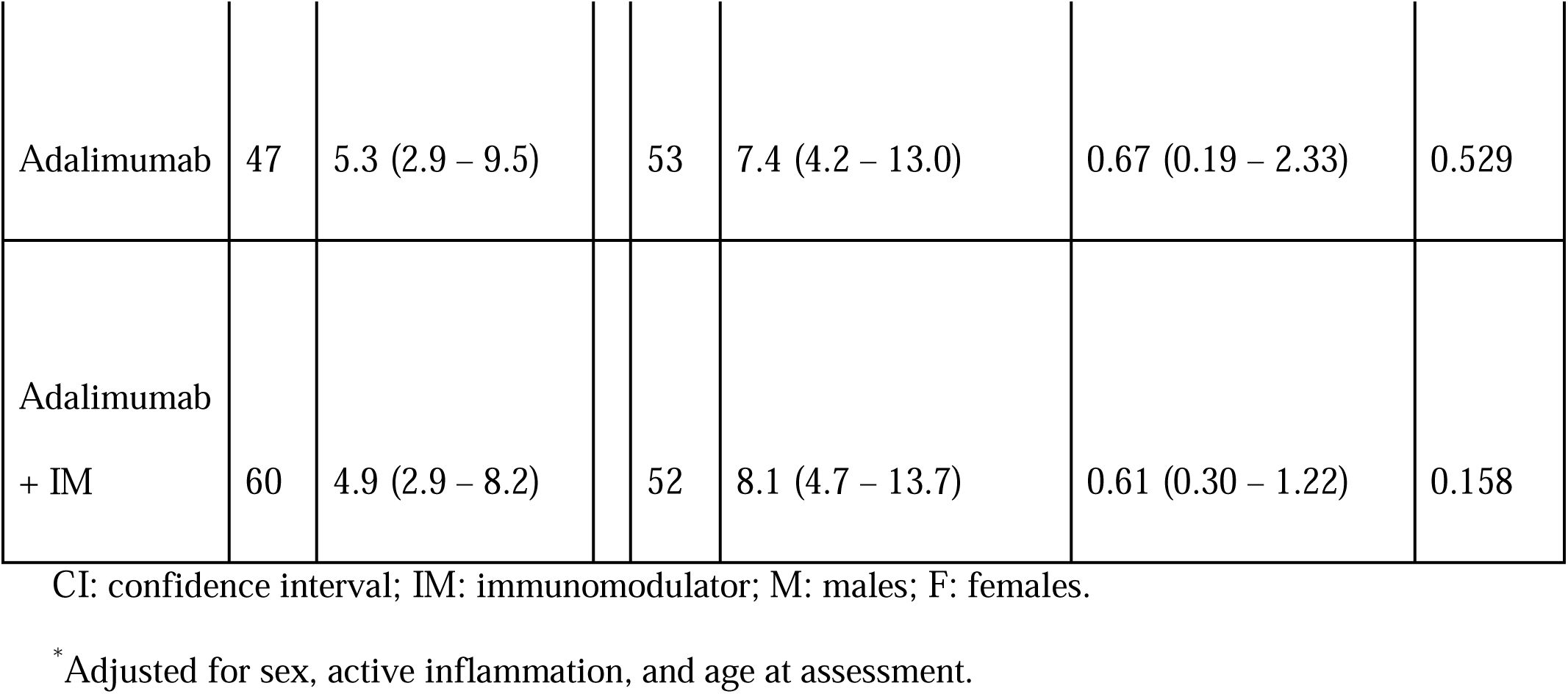
Anti-TNF serum concentrations stratified by sex according to monotherapy and combination therapy for infliximab and adalimumab use: inter-sex comparisons

Anti-drug antibody levels among patients on infliximab combination therapy were significantly lower than in patients on infliximab alone (25.7 vs. 50.8 AU/ml; aRoGM = 0.51, 95% CI: 0.28-0.91, p-value = 0.023; Supplementary Table s2). In addition, there was no difference in anti-drug antibody between patients on adalimumab combination therapy and those on adalimumab alone (12.7 vs. 12.7 AU/ml; aRoGM = 1.00, 95% CI: 0.66-1.51, p-value = 0.995; Supplementary Table s2).

In the total study sample, a negative correlation between infliximab serum concentration and anti-drug antibodies to infliximab (ATI) (spearman r = -0.388, p-value <0.001) and adalimumab serum concentration and anti-drug antibodies to adalimumab (ATA) (spearman r = - 0.415; p-value <0.001; Figure 2) was observed.

**Figure 2.**
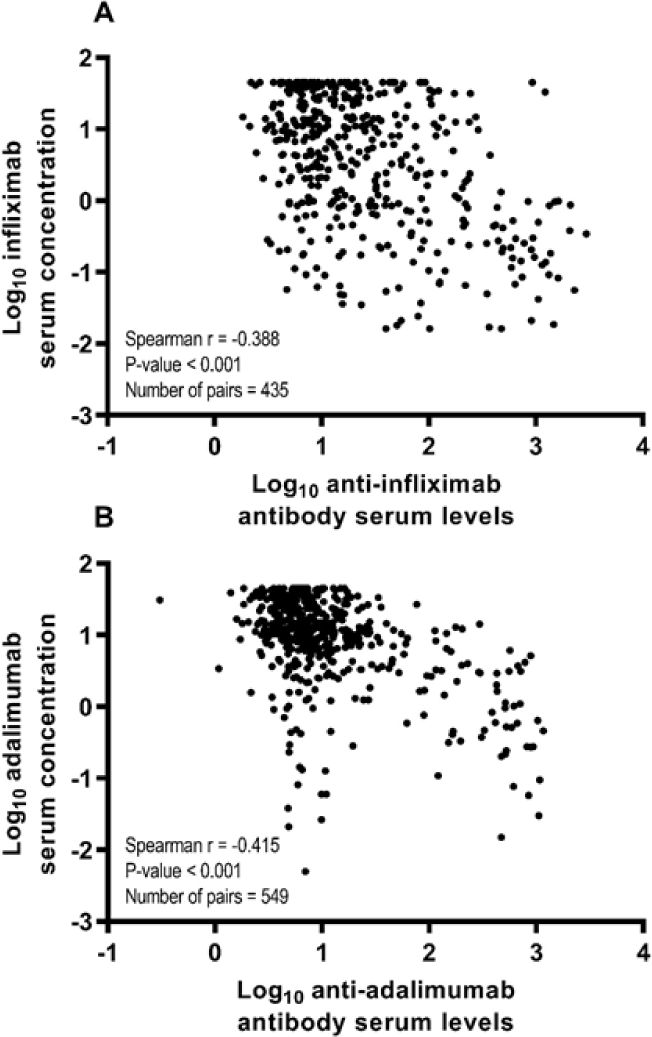
Spearman correlation coefficients (r) between anti-drug antibody levels and anti-TNF therapy serum drug concentration according to anti-TNF therapy in the total study sample. A) Spearman correlation coefficient between log_10_-transformed infliximab serum concentration and log_10_-transformed anti-infliximab antibody serum levels. B) Spearman correlation coefficient between log_10_-transformed adalimumab serum concentration and log_10_-transformed anti-adalimumab antibody serum levels.

In an additional analysis, we assessed the association of serum albumin (≥40 g/L [normal] vs. <40 g/L [abnormal]) with anti-drug antibodies to infliximab and adalimumab. Comparing subjects with normal and abnormal serum albumin levels, we found that average levels of anti-drug antibodies to infliximab (10.6 vs. 19.2 AU/ml, p-value = 0.158) and anti-drug antibodies to adalimumab (7.5 vs. 7.4 AU/ml, p-value = 0.991) to be similar across categories of serum albumin. Moreover, when including serum albumin in the multiple linear regression models, the observed associations between male sex and higher anti-drug antibodies to infliximab (aRoGM = 1.79, 95% CI: 1.02-3.11, p-value = 0.041) and male sex and lower infliximab serum drug concentrations (aRoGM = 0.73, 95% CI: 0.23-0.96, p-value = 0.046) remained statistically significant. Hence, these additional analyses indicate that albumin does not confound the reported associations.

## Discussion

In 1093 patients with inflammatory bowel disease (IBD), we evaluated the effect of patient sex and combination therapy on anti-drug antibody levels and serum drug concentrations among all patients who were on infliximab or adalimumab. The effect of the patient sex on the anti-drug antibody levels was investigated in both monotherapy and combination therapy in our subgroup analysis. The results showed that serum drug concentrations of infliximab in male patients are commonly lower than female patients. It is thought that male patients have higher clearance rates than female patients, however; the exact mechanism of this effect is still unknown ^11^. Moreover, male compared to female patients on infliximab had higher anti-drug antibody concentrations; nonetheless, there is no previously available evidence, to our knowledge, that supports the association between patient sex and presence of anti-drug antibodies. On the other hand, the presence of higher anti-drug antibody levels seems to accelerate the clearance of anti-TNF therapy, which is supported by Fasanmade et al. study that analysed two randomized-controlled trials.^22^

The available data on therapeutic drug response of patients with IBD to medications, stratified by sex, are extremely limited. A recent review by Rustgi et al. suggested the need for further investigation to the role of sex hormones on IBD, to get better therapeutic response for patients with IBD.^23^ Further studies needed to identify if there is a genetic factor behind a correlation between the patient sex and the anti-drug antibody levels. Two previous studies, by Wilson et al. and Sazonovs et al., identified an association between the genetic variant HLA-DQA1*05 and the formation of anti-drug antibodies, against both infliximab and adalimumab, in patients with Crohn’s disease (CD).^24,25^ A study by Sazonovs et al. showed that this variant increased the anti-drug antibodies formation by 2-folds, regardless of the concurrent immunomodulator use.^25^ They also concluded that to minimize the risk of therapy failure, a pretreatment genetic testing for HLA-DQA1*05 might be helpful in deciding whether to use anti-TNF, or combination therapy in IBD.

As a secondary outcome of our study, the effect of combination therapy was investigated in one inflammatory bowel disease center. The results of our study were similar to the PANTS study, which showed that immunogenicity is more common in patients with CD treated with infliximab than adalimumab, and that the concomitant use of immunomodulator was associated with lower the immunogenicity.^26^ Moreover, higher drug concentrations and remission rates were found in patients treated with infliximab combination therapy. However, these effects were not shown in patients treated with adalimumab combination therapy, this might be influenced by lower rates of immunogenicity compared to infliximab.^26^ Furthermore, our study results agree with Hazlewood et al. network meta-analysis, where the effectiveness of immunosuppressants and anti-TNF were found to be comparable, and considered the combination therapy of infliximab with azathioprine and adalimumab monotherapy to be the most effective strategies for inducing and maintaining the remission of CD.^27^ In addition, a systematic review, by Strand et al., emphasized the role of monitoring both anti-drug antibody levels and serum drug concentrations of the used anti-TNF agent.^28^ This might be potentially helpful in guiding clinicians to improve anti-TNF therapy management as well as clinical outcomes. It can also reduce risks associated with immunogenicity and help in lowering costs of therapy.^28^

Although it has been a common practice in clinical research to analyze anti-drug antibodies and drug concentrations (i.e., continuous variables) as dichotomous/categorical variables (e.g., normal/low vs. abnormal/high), such categorization is helpful in clinical practice, but has some drawbacks. Loss of information, reduced statistical power, underestimating the true variability in the data, and residual confounding are the major issues with categorization of continuous variables in clinical research.^29^ Given the previous drawbacks, we have analyzed the outcome variables (i.e., anti-drug antibodies and serum drug concentrations) as continuous variables.

Our study has several strengths. It is a nationwide multi center study that involved all centers in the country where therapeutic drug monitoring (TDM) testing are done. It is well designed with over 3 years total of all available data, of eligible patients, were collected. It also addresses a gap in knowledge and encourage future research in this area.

However, there were some limitations to our study. Being a retrospective cohort, there might be some confounders, such as those on monotherapy could have been on combination therapy previously and then were discontinued due to adequate serum drug concentration and low anti-drug antibodies. In addition, patients’ adherence to anti-TNF therapy at regular intervals could not be evaluated. Moreover, endoscopic and clinical targets were not studied; however, we controlled for objective inflammatory markers (CRP, Fcal, and steroids use). Finally, the proportion of therapeutic drug monitoring (TDM) tests that were done reactively vs proactively was not assessed.

## Conclusion

Anti-drug antibodies to infliximab (ATI) were higher in males than females whereas anti-drug antibodies to adalimumab (ATA) were similar in both sexes. In addition, male patients had lower infliximab serum drug concentrations compared to female patients while no sex differences was observed in adalimumab serum drug concentrations. Moreover, combination therapy was more effective than monotherapy in reducing ATI, but not better than monotherapy in reducing ATA. However, male and female patients on infliximab combination therapy had similar anti-drug antibody levels. Future studies are needed to assess the effect of patient sex, i.e sex hormones, on anti-TNF anti-drug antibody and serum drug concentrations in patients with inflammatory bowel disease.

## Supporting information

checklist

## Data Availability

All data available with the submitted files.

## Data Availability

Original data not available due to ethical and local legal restrictions.

## Funding

This study was funded by the Ministry of Health of Kuwait.

## Competing Interests

All authors declare no conflict of interest.

## Acknowledgment

None.

## Preprint

A preprint has previously been published.^30^

## Notes

### Competing Interest Statement

The authors have declared no competing interest.

### Funding Statement

Funded by Ministry of Health

### Author Declarations

The study was reviewed and approved by the standing committee for coordination of health and medical research at the Ministry of Health in Kuwait (IRB 2020/1410).

