## supplementary documents for "Relationship Between Patient Sex and Serum Tumor Necrosis Factor Antagonist Drug and Anti-Drug Antibody Concentrations in Inflammatory Bowel Disease; A Nationwide Cohort Study"

### Supplementary Material

**Table s1.** Anti-TNF serum concentrations according to monotherapy and combination therapy: comparing "infliximab + immunomodulator to infliximab" and "adalimumab + immunomodulator to adalimumab" in the subsample†

| | | Anti-TNF **serum concentration, ug/ml** | | |
|---|---|---|---|---|
| **Drug** | **n** | **Geometric mean (95% CI)** | **Ratio of geometric means* (95% CI)** | **P-value** |
| Infliximab | 33 | 1.1 (0.6 – 2.2) | 1.00 (Reference) | – |
| Infliximab + IM | 114 | 1.8 (1.2 – 2.7) | 1.57 (0.77 – 3.20) | 0.217 |
| Adalimumab | 93 | 6.3 (4.0 – 9.7) | 1.00 (Reference) | – |
| Adalimumab + IM | 108 | 6.3 (4.2 – 9.3) | 1.00 (0.60 – 1.67) | 0.987 |

CI: confidence interval; IM: immunomodulator.

* Adjusted for sex, active inflammation, and age at assessment.

† Includes patients with information on combination therapy use and active inflammation.

**Table s2.** Anti-drug antibody serum levels according to monotherapy and combination therapy: comparing "infliximab + immunomodulator to infliximab" and "adalimumab + immunomodulator to adalimumab" in the subsample†

| | | Anti-drug antibody serum levels, AU/ml | | |
|---|---|---|---|---|
| **Drug** | **n** | **Geometric mean (95% CI)** | **Ratio of geometric means* (95% CI)** | **P-value** |
| Infliximab | 33 | 50.8 (29.3 – 88.1) | 1.00 (Reference) | – |
| Infliximab + IM | 114 | 25.7 (18.3 – 36.2) | 0.51 (0.28 – 0.91) | 0.023 |
| Adalimumab | 100 | 12.7 (8.9 – 18.1) | 1.00 (Reference) | – |
| Adalimumab + IM | 112 | 12.7 (9.3 – 17.4) | 1.00 (0.66 – 1.51) | 0.995 |

CI: confidence interval; IM: immunomodulator.

* Adjusted for sex, active inflammation, and age at assessment.

† Includes patients with information on combination therapy use and active inflammation.
